# Post Intensive Care Syndrome (PICS) in COVID-19 ARDS Survivors: A 6-Month Longitudinal Study from South India

**DOI:** 10.1101/2025.07.07.25330923

**Authors:** Azhakath Suresh Abhay, V. Parvathy Narayanan, T. Gopinathan, Vijay Sundar Singh

**Author notes:** Email id, Address: Thattakam 144, Sree Vihar, Angadippuram, Malappuram district, Kerala, India, Pin-679321. Co- Authors (V. Parvathy Narayanan, T. Gopinathan, Vijay Sundar Singh). Email id, Email id, Email id.

## Abstract

**Background:** Post-Intensive Care Syndrome (PICS) includes cognitive, psychological, and physical impairments following critical illness. The long-term impact of COVID-19-related ARDS on PICS domains remains under-explored, particularly in resource-limited settings.

**Objective:** To assess the prevalence and trajectory of cognitive, mental, and physical impairments among COVID-19 ARDS survivors at ICU discharge and at 6 months, and to explore associated risk factors.

**Methods:** This was an observational cohort study of 30 mechanically ventilated COVID-19 patients admitted to a tertiary ICU in South India during the second wave (Delta variant). Patients were assessed at ICU discharge (or first follow-up) and at 6 months using the Montreal Cognitive Assessment (MoCA), SF-36 health survey, modified MRC dyspnea scale, 6-minute walk test (6MWT), and physical examination. Risk factors were analyzed using multivariable linear regression.

**Results:** At discharge, mild cognitive impairment was prevalent (MoCA: 25.17±3.63), with significant improvement at 6 months (27.07±2.72, p<0.001). SF-36 domains showed persistent deficits in emotional well-being (36.31→50.83), fatigue (33.28→45.91), and pain (51.38→71.47) (all p<0.01). Functional capacity improved on 6MWT, with >350m walked increasing from 23% to 53%. Risk factors included steroid duration, SOFA score, antifungal exposure, and fasting hypoglycemia. Other parameters like Muscle wasting, dyspnea, and gastrointestinal symptoms also showed partial recovery.

**Conclusion:** COVID-19 ARDS survivors experience significant but partially reversible PICS across multiple domains. Structured post-ICU rehabilitation and early identification of modifiable risk factors may improve recovery trajectories. Findings highlight the need for integrated post-ICU care pathways in similar settings.

## INTRODUCTION

There is a growing population of survivors of critical illness due to advances in critical care medicine. Many survivors experience impairment in cognition, mental health, and physical function, known as post-intensive care syndrome (PICS). There is no established definition for post-intensive care syndrome (PICS), the epidemiology has not been studied in detail in developing countries and the exact prevalence is unknown. Most clinicians agree that PICS constitutes new or worsening function in one or more of the following domains after critical illness: Cognitive function, Mental function, Physical function ^(1)^. These impairments persist long after the intensive care unit (ICU) hospitalization up to 5 to 15 years.

One-half or more of ICU patients probably will suffer from some component of PICS (cognitive, psychiatric, physical dysfunction)^(1,3–7)^. Risk factors for the post-intensive care syndrome (PICS) have not been clearly defined and vary depending on the component of PICS that is studied^(8)^. Commonly cited risk factors can be broadly categorized into pre-existing factors (e.g. neuromuscular disorders, dementia, psychiatric illness, comorbid conditions) and intensive care unit (ICU)-specific factors (e.g. mechanical ventilation, acute delirium, sepsis, acute respiratory distress syndrome)^(9)^. We still are not aware if critical illness accelerates preexisting neuropsychological and functional decline or if it unmasks underlying illness.

Acute respiratory distress syndrome (ARDS) is a clinical syndrome of diffuse lung inflammation and oedema that causes acute respiratory failure^(10)^. They may spend weeks in the ICU and are vulnerable to the cumulative insults of ICU care in the context of catabolism and systemic inflammation^(11–13)^. New disability is found in one out of three survivors and the incidence and severity of disability is similar for survivors of COVID-19-related critical illness^(14)^.

The coronavirus disease 2019 (COVID-19) pandemic has resulted in a growing population of individuals recovering from severe ARDS. Previous studies suggest that these patients may experience a wide range of symptoms after recovery from acute illness, referred to by several terms including “long COVID,” “post-COVID conditions,” and “postacute sequelae of SARS-CoV-2 infection.” Some aspects of this recovery may be unique to COVID-19, but many appear to be similar to recovery from other viral illnesses, critical illness, and/or sepsis^(15–20)^. PICS may be considered as a long term manifestation of ‘Long Covid’ seen in patients admitted to ICU though it may also be seen in other patients with ARDS^(21–24)^. The multiple variants and the mutational ability of the virus limits generalizability. The delta variant is the focus of this study as it was the predominant variant during the study period.

This research work analyses PICS in Covid-19 patients with severe ARDS, who have been subjected to multiple new, less used treatment modalities, by combined assessment of their cognitive, mental and physical domains. Risk factors for the long term consequences of Sars CoV-2 ARDS were also isolated.

## MATERIALS AND METHODS

***Study setting:*** Follow up of patients with Sars Cov-2 infection ARDS who underwent mechanical ventilation at time of discharge and 6 months.

***Study location***: Kovai Medical Center and Hospital, 99, Avinashi Road, Coimbatore, Tamil Nadu – 641014.

***Study Design :*** Observational cohort study with retrospective identification and prospective follow-up

***Study Duration :***From Date of ethics committee approval to February 2023

**The retrospective nature of study was because of delay in ethics committee meeting and subsequent approval in view of Covid-19 crisis.

### Inclusion criteria

1. RTPCR positive for Covid-19. RTPCR was done for every patient in the study. It had the capability to detect the RNA dependent RNA polymerase (RdRP), Envelope (E), Nucleocapsid (N), Open reading frame (Orf1ab) genes. The alpha and omicron variants had S gene dropout and hence can be differentiated from Delta variant. However whole genome sequencing wasn’t done.
2. Other causes for ARDS unlikely
3. Covid-19 survivors in the 2^nd^ wave from April to September 2021 attributed to Delta Variant.
4. Covid-19 survivors who have undergone mechanical ventilation in our ICU for 4 days or more.
5. Covid-19 survivors who had P/F ratios 200 or less.
6. Covid-19 survivors who have given consent.

### Exclusion criteria

1. Patients with Stroke or Trauma at the time of admission.
2. Patients with chronic lung disease at the time of admission
3. Patients who were immobile at the time of ICU admission
4. Patients not giving consent

### SAMPLE SIZE

At the time the study was undertaken, there were very few studies internationally and no studies in India about this topic. I have done the study in patients who fulfill the inclusion and exclusion criteria and those who have given consent. We used convenience sampling for our study and hence we acknowledge the limitations that may arise due to underpowering. 300 patients admitted to our ICU with Covid-19 were screened and 62 were selected as per inclusion criteria, out of 62 another 25 were lost to follow up and another 7 patients were excluded in view of other acute illness during study period. Hence strict follow up was obtained for 30 patients and this convenient sample size was taken for this study.

### METHODS

All Sars CoV-2 (Covid -19) survivors after ICU admission in our Centre, during the second wave of its spread in India (approximately from April 2021 to September 2021) underwent interviews aimed to assess cognitive dysfunction, mental health status and detailed physical examination to assess any impairment at the time of their discharge or at 1^st^ follow up visit. This process was repeated 6 months later. The interviews were done face to face by trained clinicians as much as feasible, by video call or over phone considering the pandemic situation at that time.

The tools used for assessing Cognitive impairment were the MOCA score, mental health abnormality as per Medical Outcomes Study Short Form (SF)-36 questionnaire, physical impairment by clinical examination, single breath count test and review of their 6 minute walk test scores. The tools were explained in local languages whenever English could not be understood.

### BASELINE CHARACTERISTICS

For data collection a proforma was designed which incorporated the patients demographic details, gender, age, APACHE 2 score at admission, SOFA score at admission, details of treatment received, duration of ICU stay, duration of hospital stay, duration of mechanical ventilation, in hospital sepsis, AKI, any Surgery undergone (MOCA score; SF – 36 questionnaire single breath count tests, mMRC dyspnea scale, 6 minute walk test details, presence or absence of edema, muscle wasting, gastrointestinal disturbances and other disorders of smell, taste or vision.

A MoCA <26 was the threshold for cognitive impairment, SF 36 components had scores of 1-100 for each domains assessed, where scores closer to 100 indicated improvement and scores closer to 1 indicated increased severity of impairment.

The degree of impairment in all the domains was ascertained and any risk factors for the same among baseline characteristics will be sought.

### STATISTICAL METHODOLOGY

Statistical analysis was performed using SPSS 26 software. Tests were chosen according to sample size and data distribution. Descriptive statistics were used along with chi squared tests, chi squared values; paired T tests to compare the cognitive, mental, physical domains. These were then subjected to multivariable linear regression analysis to ascertain risk factors.

## OBSERVATION AND RESULTS

### Baseline characteristics

Out of a total of 30 patients included in the study, 73.33% were male. Of the males who survived, 11 resumed work by 6 months. One patient died after discharge before 6 month follow up. Two patients were pregnant at the time of diagnosis. Two patients in the study population required ECMO support. Mean Age at presentation was 50.57. The mean duration of hospital stay was 38.47 days. The mean duration of ICU stay was 17.73 days. The mean duration of mechanical ventilation was 20.30 days. The mean APACHE 2 and SOFA scores in first 24 hours was 11.07 and 5.20 respectively. **Table 1**

**Table 1:** Baseline characteristics of study population.

| Variables | Mean | Range | Minimum | Maximum | Std. Deviation | Variance |
| --- | --- | --- | --- | --- | --- | --- |
| **Age** | 50.57 | 58 | 19 | 77 | 16.488 | 271.840 |
| **Days of Hospital Stay** | 38.47 | 80 | 10 | 90 | 21.919 | 480.464 |
| **Days of ICU stay** | 17.97 | 44 | 1 | 45 | 12.527 | 156.930 |
| **Days of Mechanical Ventilation** | 20.30 | 60 | 4 | 60 | 16.472 | 271.321 |
| **APACHE 2 score at admission** | 11.07 | 24 | 5 | 24 | 5.601 | 31.375 |
| **SOFA score at admission** | 5.20 | 12 | 2 | 12 | 2.683 | 7.200 |

Among the study subjects 23.33% received 4 weeks of steroids, 20% received 3 weeks of steroids, 13.33% received 1 week of steroids; 10% received 2 weeks, another 10%, 8 weeks and yet another 10% received more than 8 weeks of steroids. 6.66% received 6 weeks of steroids; 3.33% received 5 weeks and another 3.33% received 7 weeks of steroids respectively. **Figure 1** Most of the patients 60% had CT severity score between 16-20 indicating severe disease. 16.67% patients had severity scores of 11-15 indicating moderate disease. Mild disease was seen with CTSS less than 11 with 6.67% patients having scores between 6-10 and 16.67% patients having scores between 1-5. **Figure 2**

**Figure 1:**
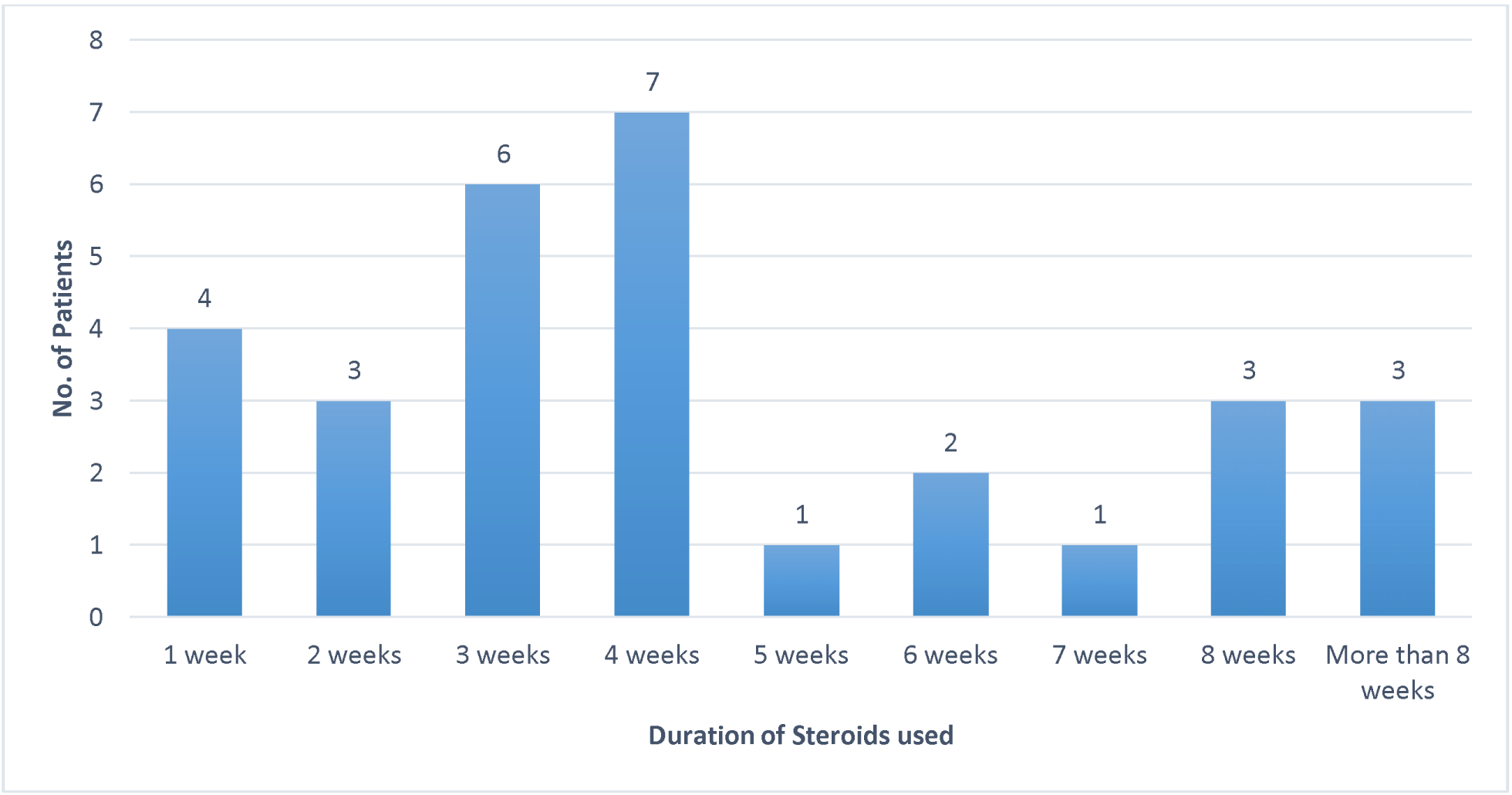
Bar graph showing duration of steroids.

**Figure 2:**
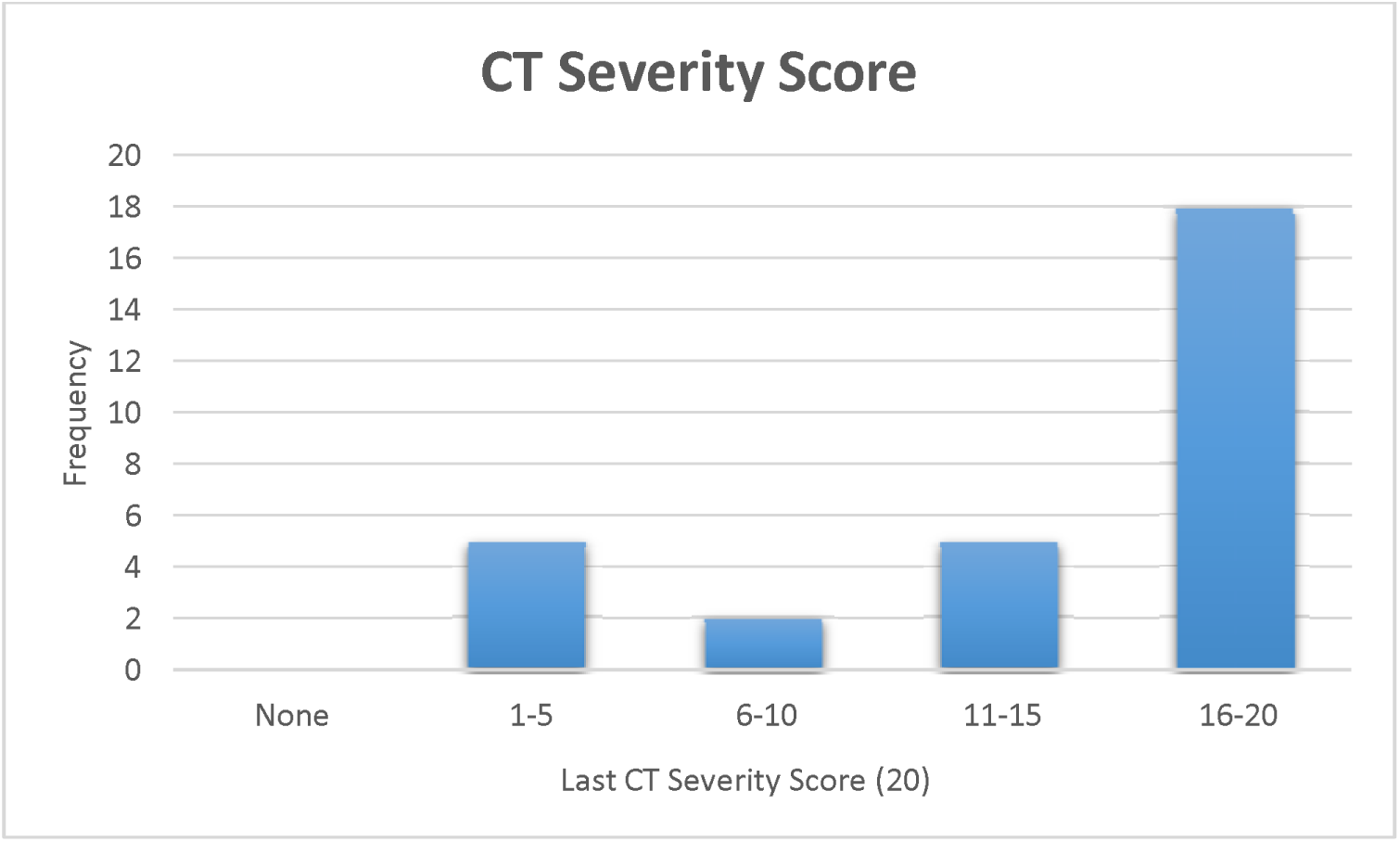
Bar graph showing HRCT Severity score.

## PICS EVALUATION

### Cognition

At 0 months all the patients had a mean MOCA score of 25.17 and on follow up at 6 months, they had a mean score of 27.07 with a paired t test showing significance. **Figure 3**

**Figure 3:**
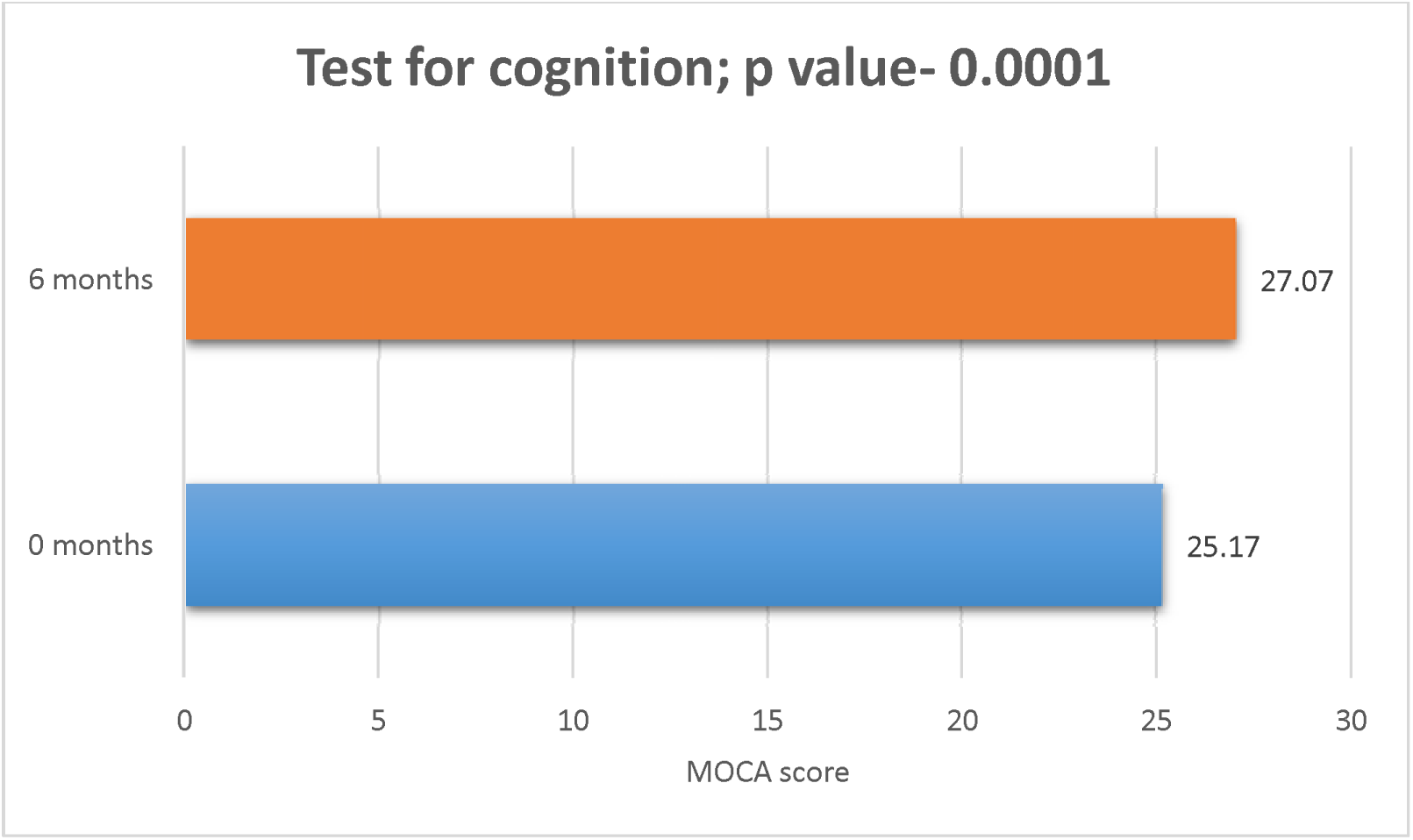
Bar graph showing MOCA scores at 0 and 6 months.

### Mental Health

The mental health was assessed as per medical outcomes study short form (SF 36) domains as shown in **Table 2**.

**Table 2:**
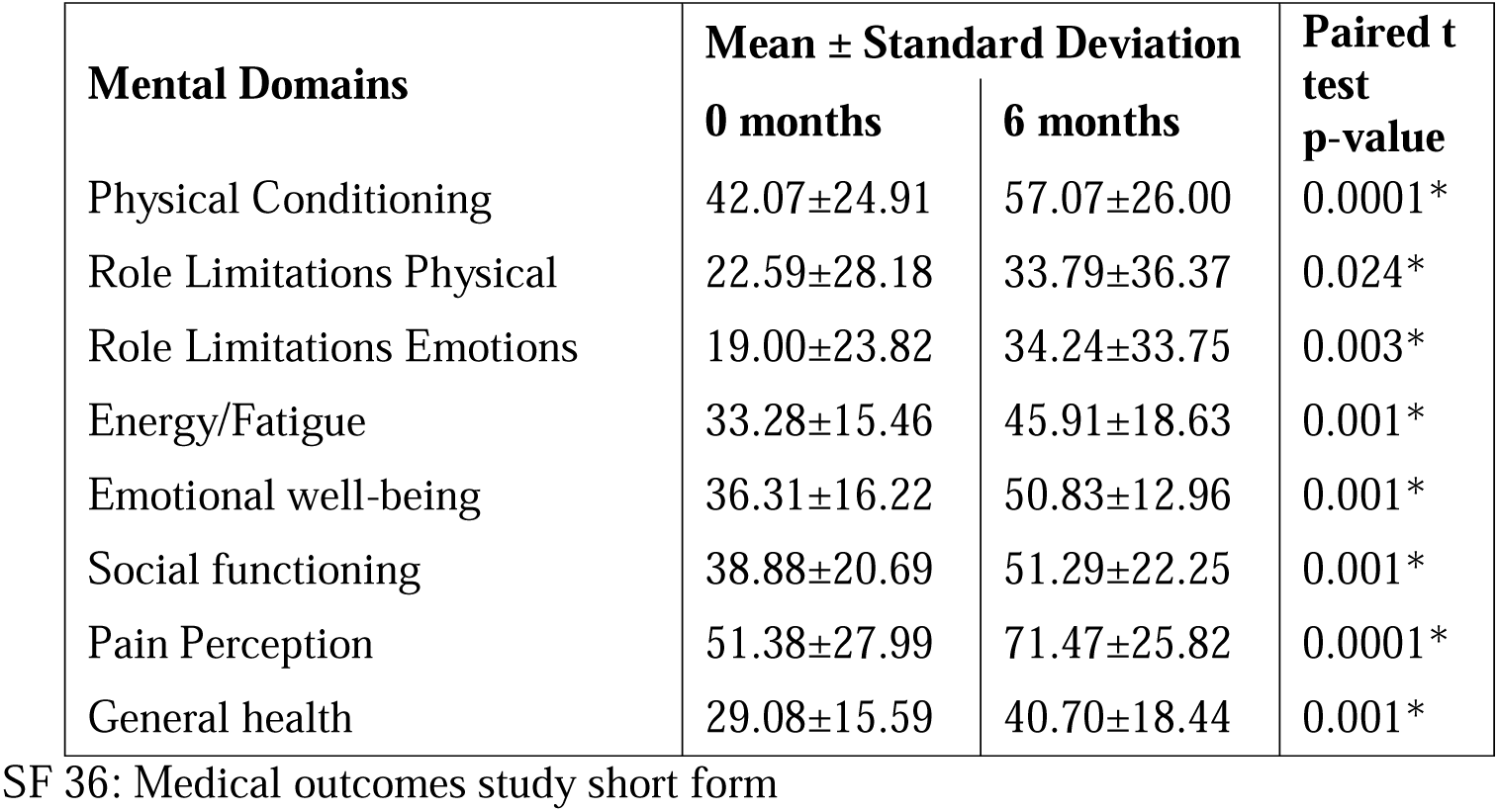
Medical outcomes study short form (SF 36) domains at 0 months and 6 months.

| **Mental Domains** | **Mean ± Standard Deviation** |  | **Paired t test p-value** |
| --- | --- | --- | --- |
|  | **0 months** | **6 months** |  |
| Physical Conditioning | 42.07±24.91 | 57.07±26.00 | 0.0001* |
| Role Limitations Physical | 22.59±28.18 | 33.79±36.37 | 0.024* |
| Role Limitations Emotions | 19.00±23.82 | 34.24±33.75 | 0.003* |
| Energy/Fatigue | 33.28±15.46 | 45.91±18.63 | 0.001* |
| Emotional well-being | 36.31±16.22 | 50.83±12.96 | 0.001* |
| Social functioning | 38.88±20.69 | 51.29±22.25 | 0.001* |
| Pain Perception | 51.38±27.99 | 71.47±25.82 | 0.0001* |
| General health | 29.08±15.59 | 40.70±18.44 | 0.001* |

In the domain of physical conditioning the mean score of 42.07 at 0 months increased to 57.07 after 6 months. Role limitations due to physical condition had a score of 22.59 at 0 months which increased to 33.79 at 6 months. Role limitations due to emotions increased from 19 at 0 months to 34.24 at 6 months. Energy levels increased from 33.28 at 0 months to 45.91 at 6 months. Emotional well being increased from 36.31 at 0 months to 50.83 at 6 months. Social Functioning at 0 months had a score of 38.88 which increased to 51.29 at 6 months. The domain of pain perception at 0 months was 51.38 which increased to 71.47 at 6 months. Overall General health increased from 29.08 at 0 months to 40.7 at 6 months. At 6 months the domains of role limitations due to emotions, physical condition and pain perception showed a significant increase at follow up while all other domains showed slight improvement though not full recovery.

### Physical Examination

#### Dyspnea scale

Grade 1 Dyspnea was seen in 20% patients at 0 months and in 10% patients at 6 month follow up. 36.67% patients had grade 2 Dyspnea on the mMRC Dyspnea Scale at 0 months and 40% patients had grade 2 dyspnea at 6 months. At 0 months, 20% patients had Grade 3 dyspnea on the mMRC Dyspnea scale and 13.33% had grade 3 dyspnea on the mMRC Dyspnea scale at 6 months. At 0 months, 10% patients had grade 4 dyspnea on the mMRC Dyspnea scale while none had grade 4 dyspnea at 6 months. **Table 3**

**Table 3:** mMRC Dyspnea Scale at 0 months and 6 months.

| **mMRC Dyspnea** | **0 months** |  | **6 months** |  |
| --- | --- | --- | --- | --- |
|  | **No. of Patients** | **Percent** | **No. of Patients** | **Percent** |
| Grade 0 | 4 | 13.33% | 10 | 33.33% |
| Grade 1 | 6 | 20.00% | 3 | 10.00% |
| Grade 2 | 11 | 36.67% | 12 | 40.00% |
| Grade 3 | 6 | 20.00% | 4 | 13.33% |
| Grade 4 | 3 | 10.00% | 0 | 0.00% |
| Total | 30 | 100.00% | 29 | 96.67% |
| Chi-square = 27.938, p-value = 0.006* |  |  |  |  |

The results proved to be statistically significant. **Figure 4**

**Figure 4:**
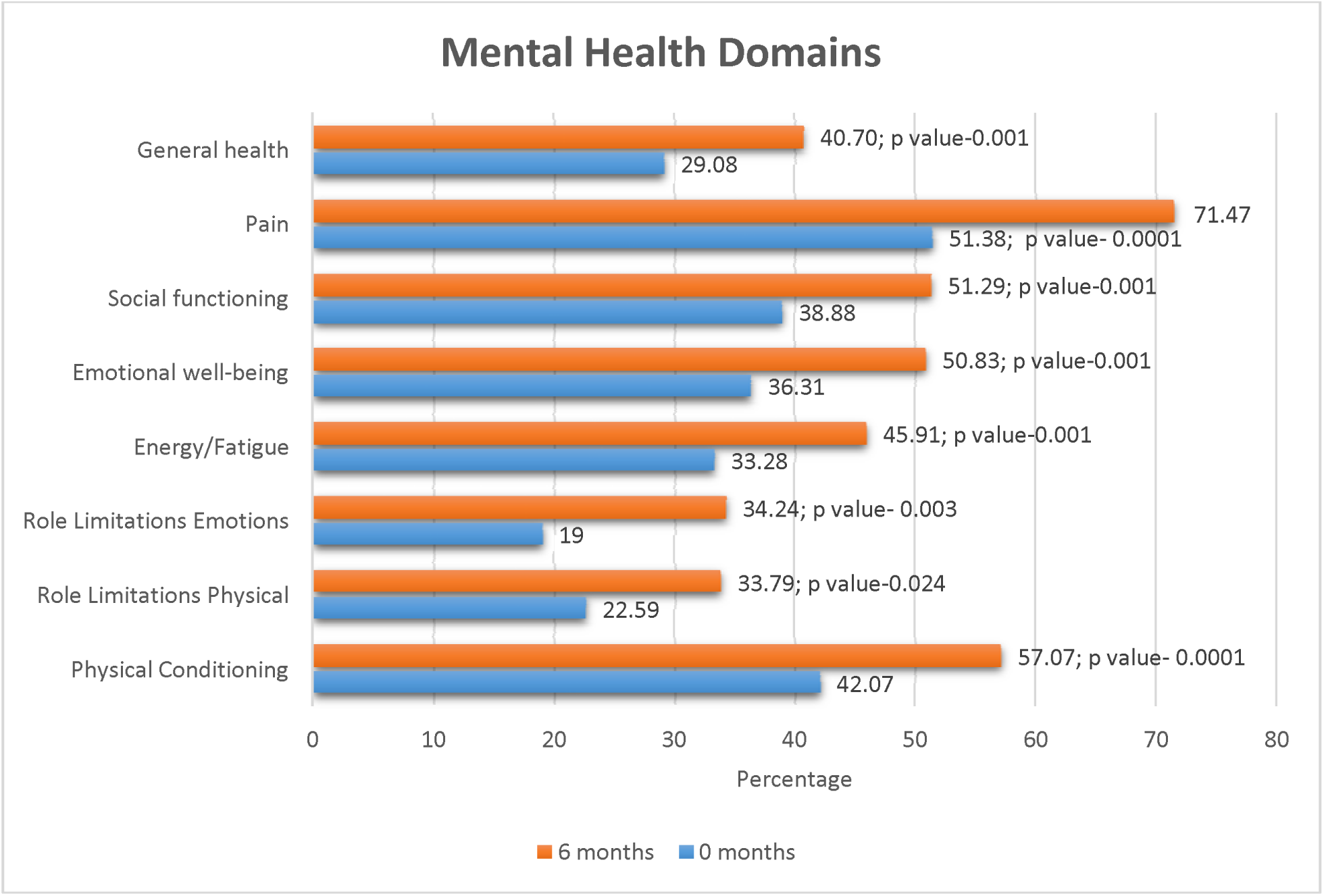
Medical outcomes study short form (SF 36) domains at 0 months and 6 months.

#### 6 Minute Walk Test (6 MWT)

At 0 months, 20% patients had a 6 Minute Walk Test of (6MWT) of <149m, while 3.33% patient had a 6MWT of <149m at 6 months. A normal 6MWT of > 350m was seen in 23.33% at 0 months and 53.33% at 6 months respectively. **Figure 5**

**Figure 5:**
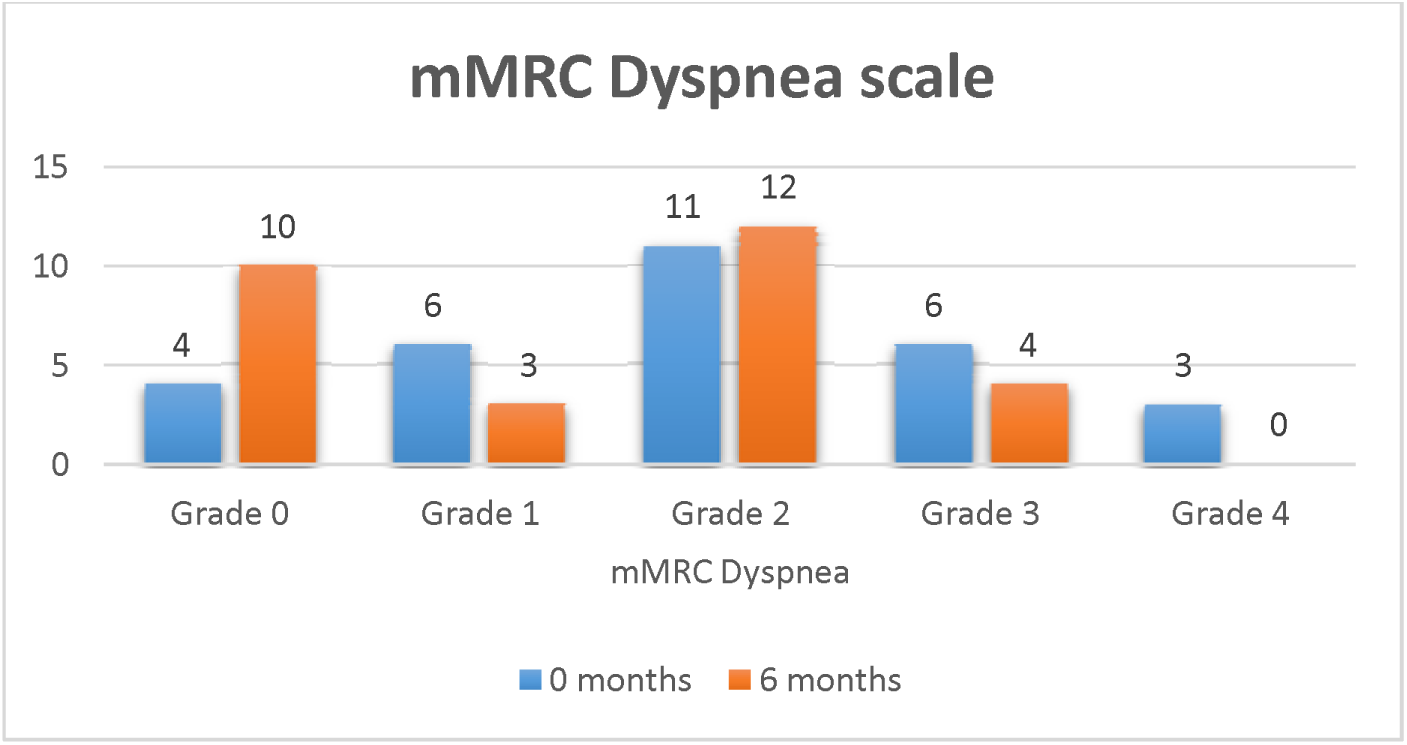
Bar Graph showing mMRC Dyspnea Scale at 0 months and 6 months.

**Figure 6:**
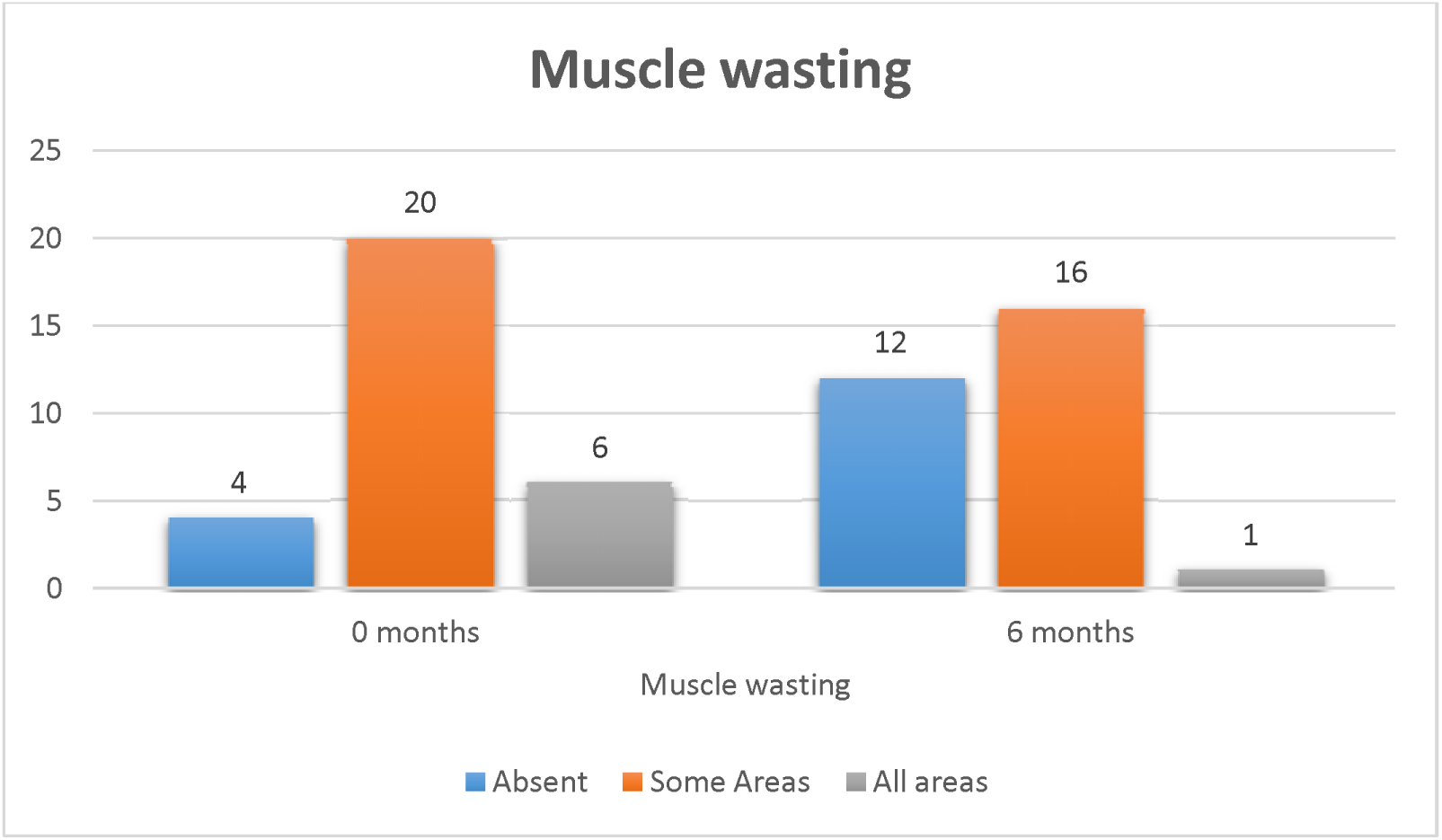
Bar Graph showing Muscle wasting at 0 months and 6 months.

**Figure 7:**
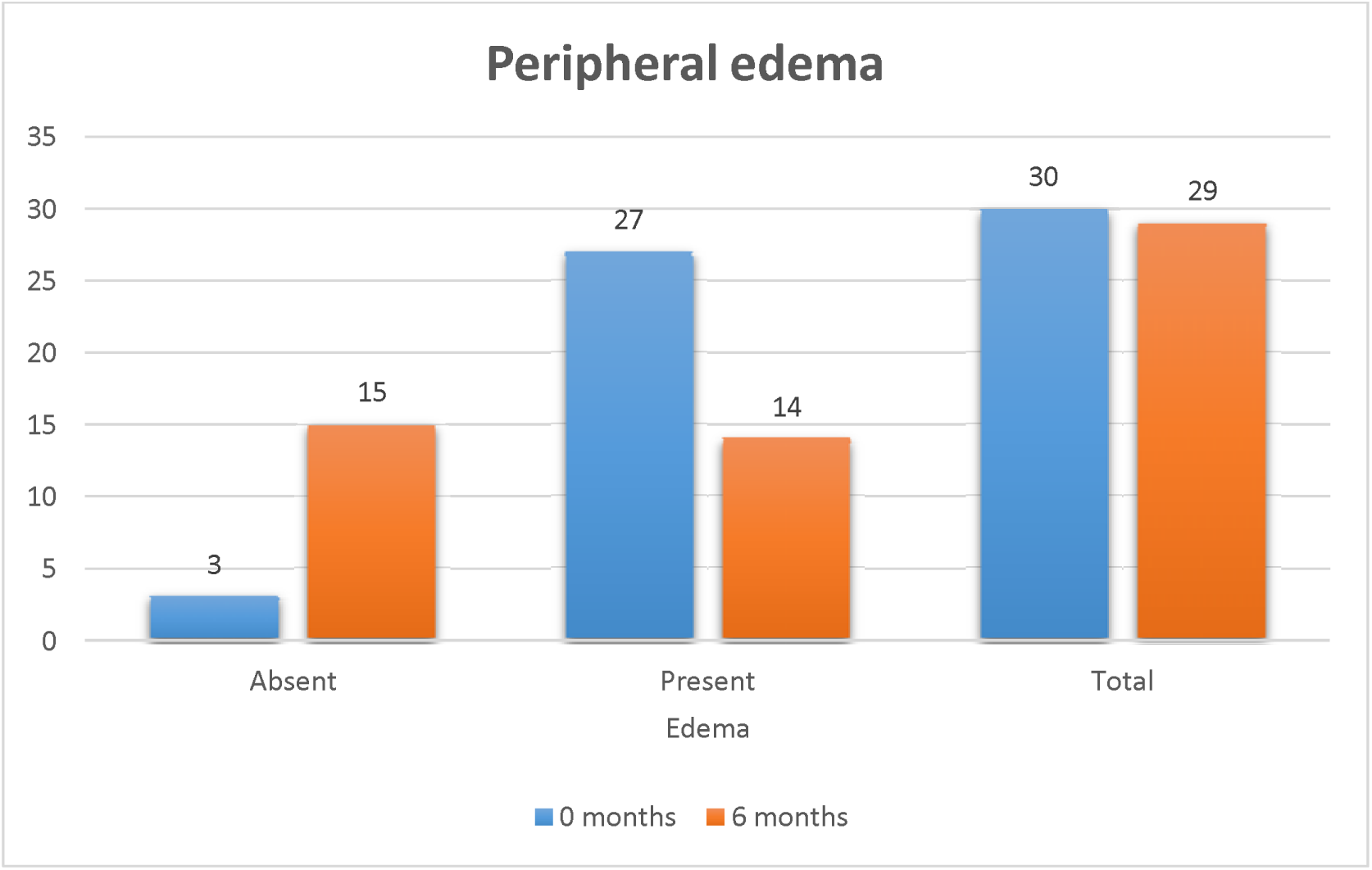
Bar Graph showing Edema at 0 months and 6 months.

**Figure 8:**
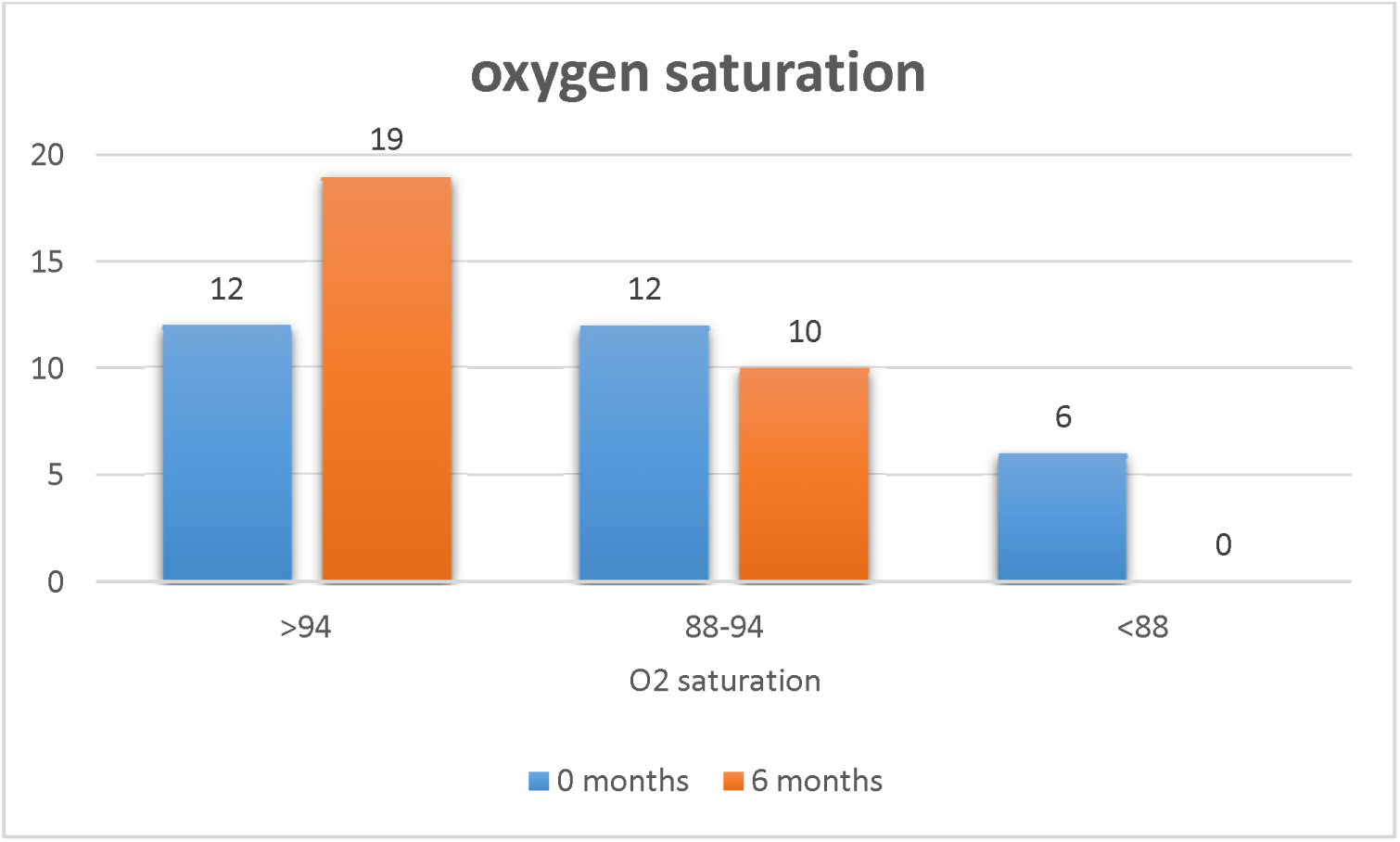
Bar graph showing Oxygen saturation at 0 months and 6 months.

**Figure 9:**
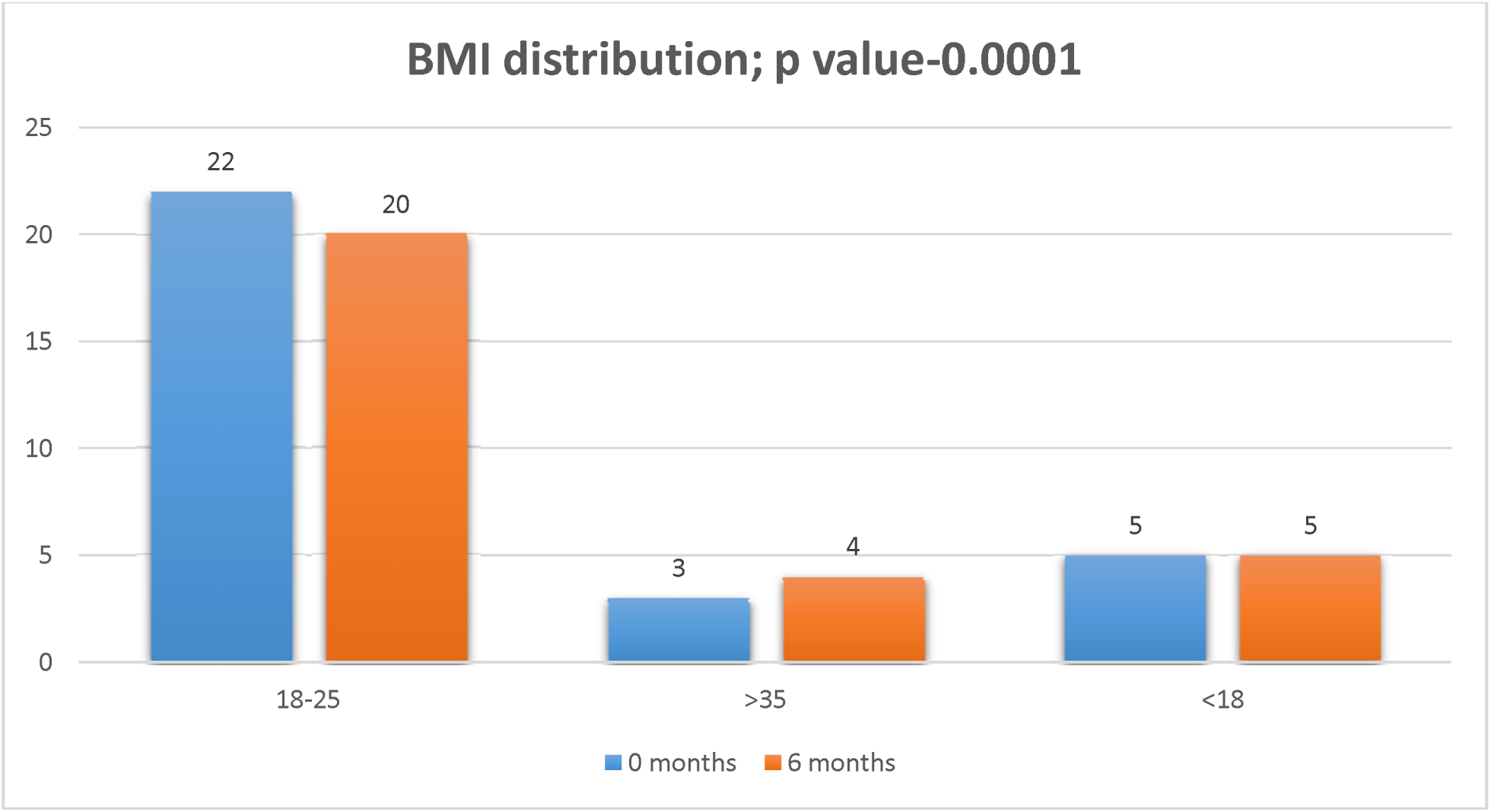
Bar graph showing BMI at 0 and 6 months.

**Figure 10:**
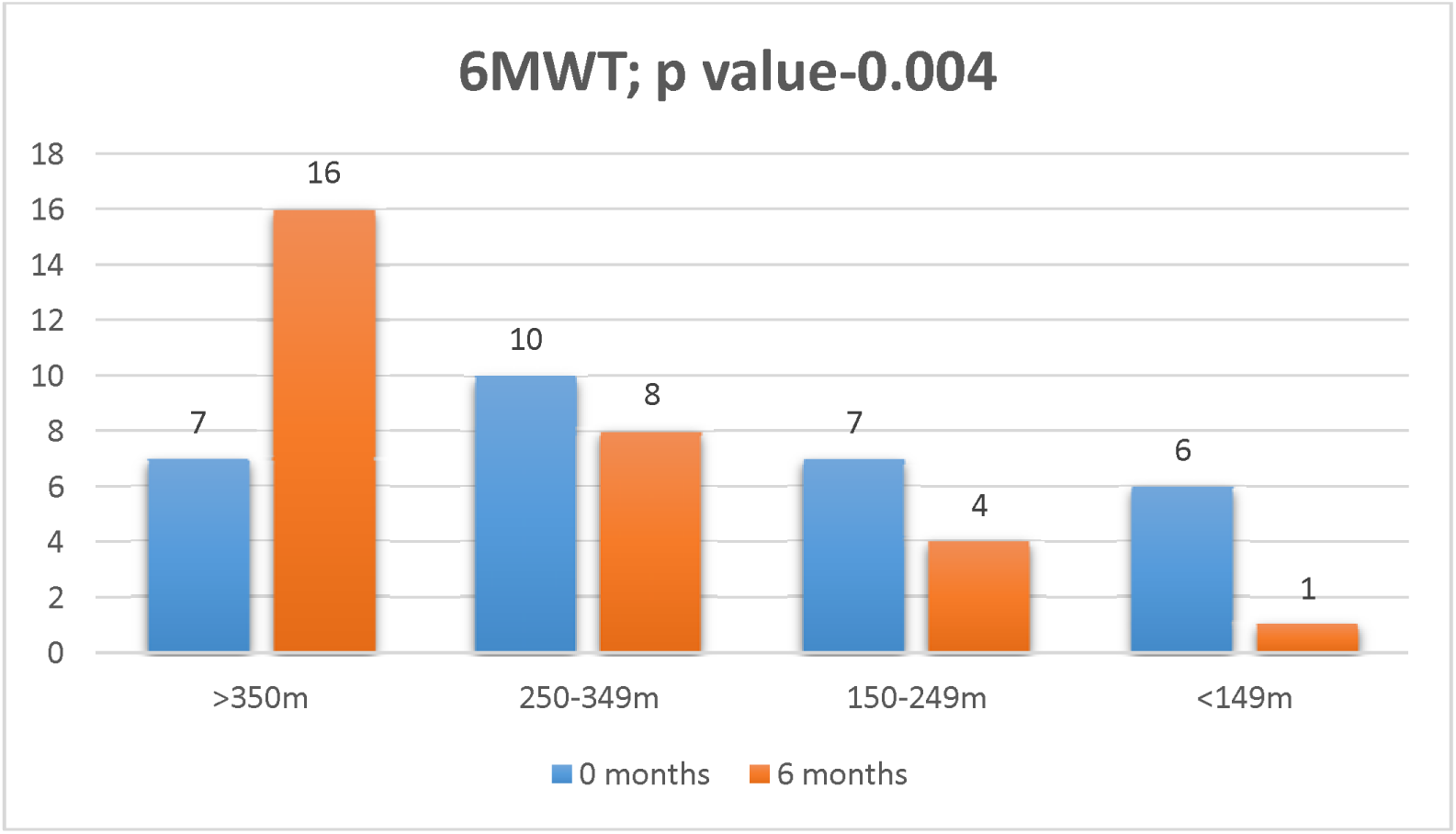
Bar Graph showing 6-minute walk test (6MWT) at 0 months and 6 months.

The statistical analysis of 6MWT over the two periods turned out to be significant with a favorable chi square test and p value. **Table 4**

**Table 4:**
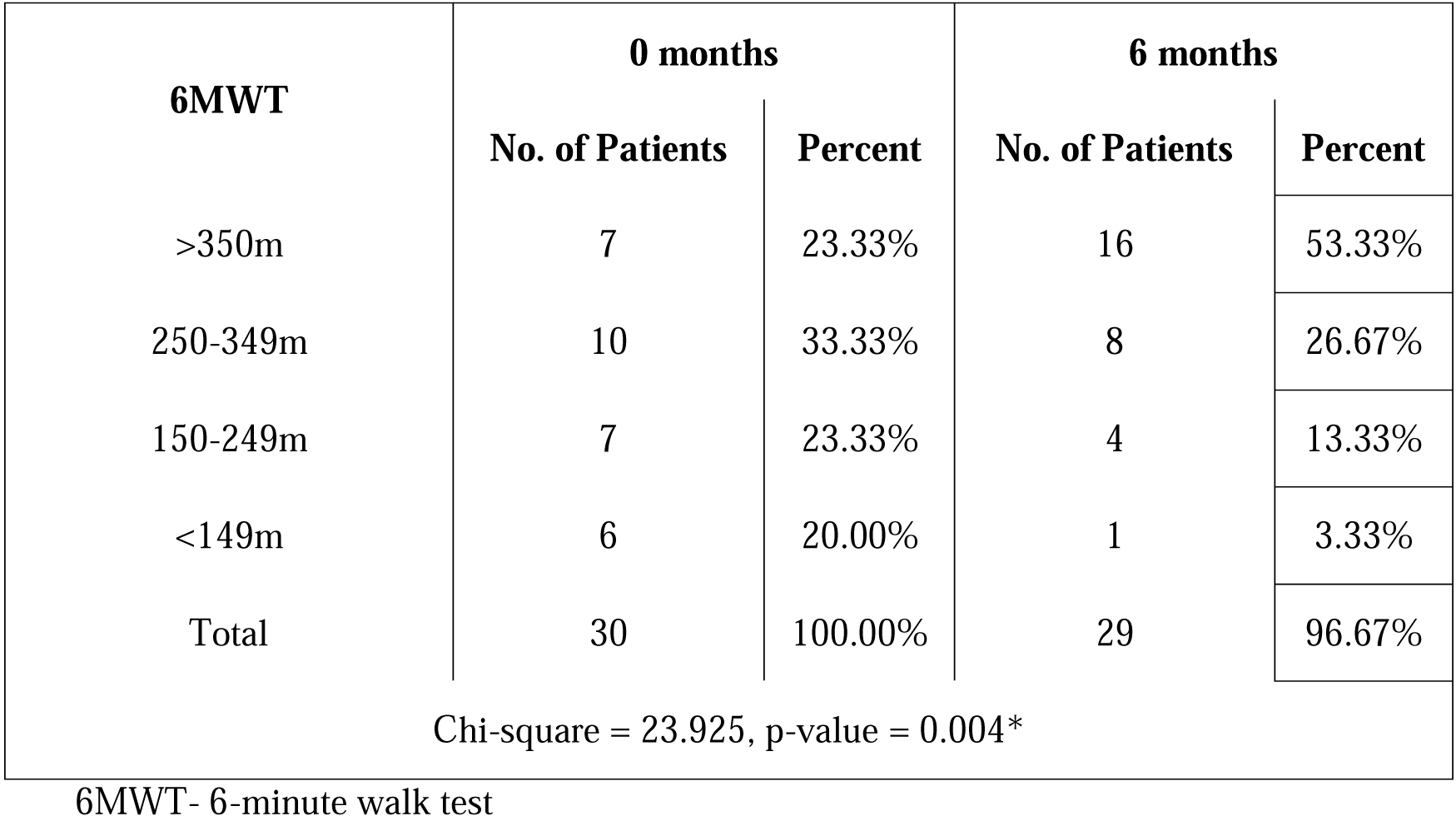
6-minute walk test (6MWT) at 0 months and 6 months.

| 6MWT | 0 months |  | 6 months |  |
| --- | --- | --- | --- | --- |
|  | No. of Patients | Percent | No. of Patients | Percent |
| >350m | 7 | 23.33% | 16 | 53.33% |
| 250-349m | 10 | 33.33% | 8 | 26.67% |
| 150-249m | 7 | 23.33% | 4 | 13.33% |
| <149m | 6 | 20.00% | 1 | 3.33% |
| Total | 30 | 100.00% | 29 | 96.67% |
| Chi-square = 23.925, p-value = 0.004* |  |  |  |  |

#### Risk factors for Post Intensive Care Syndrome

When assessed for risk factors for all the domains of PICS at 0 and 6 months, none of the selected baseline characteristics showed relevance on the development of PICS based on a non-significant p value. **Table 5**

**Table 5:** Risk factors for Post Intensive Care Syndrome components among baseline characteristics.

| Risk Factors | 0 months |  | 6 months |  |
| --- | --- | --- | --- | --- |
|  | P Value | Power | P Value | Power |
| **Age** | 0.251 | 0.163 | 0.781 | 0.063 |
| **Gender** | 0.477 | 0.092 | 0.586 | 0.078 |
| **Antiviral use** | 0.457 | 0.095 | 0.422 | 0.102 |
| **Antibiotic use** | 0.401 | 0.106 | 0.867 | 0.059 |
| **Antifungal use** | 0.249 | 0.164 | 0.335 | 0.125 |
| **Use of Immunomodulators** | 0.804 | 0.062 | 0.935 | 0.056 |
| **Duration of steroids** | 0.534 | 0.084 | 0.458 | 0.095 |
| **CT Severity score** | 0.853 | 0.060 | 0.309 | 0.134 |
| **Days of Hospital Stay** | 0.813 | 0.062 | 0.559 | 0.081 |
| **Days of ICU stay** | 0.881 | 0.058 | 0.164 | 0.243 |
| **Days of Mechanical Ventilation** | 0.742 | 0.066 | 0.291 | 0.141 |
| **APACHE2** | 0.862 | 0.059 | 0.906 | 0.057 |
| **SOFA** | 0.318 | 0.130 | 0.826 | 0.061 |
| **Fasting Hypoglycemia** | 0.707 | 0.068 | 0.268 | 0.153 |
| **Septic Shock** | 0.378 | 0.112 | 0.339 | 0.123 |
| **AKI stage** | 0.541 | 0.083 | 0.544 | 0.082 |
| **Surgery Undergone** | 0.862 | 0.059 | 0.982 | 0.053 |

### At 0 months

Post hoc multivariate analysis using generalized linear regression showed that at 0 months, GIT symptoms had Female Gender, antibiotic use and SOFA score at admission as risk factors. Muscle wasting had duration of hospital stay as a risk factor. Edema had duration of steroid use as risk factor. Oxygen saturation had antifungal use and duration of steroid use as risk factors. 6 Minute walk test had Fasting hypoglycemia as risk factor. **Table 6**

**Table 6:**
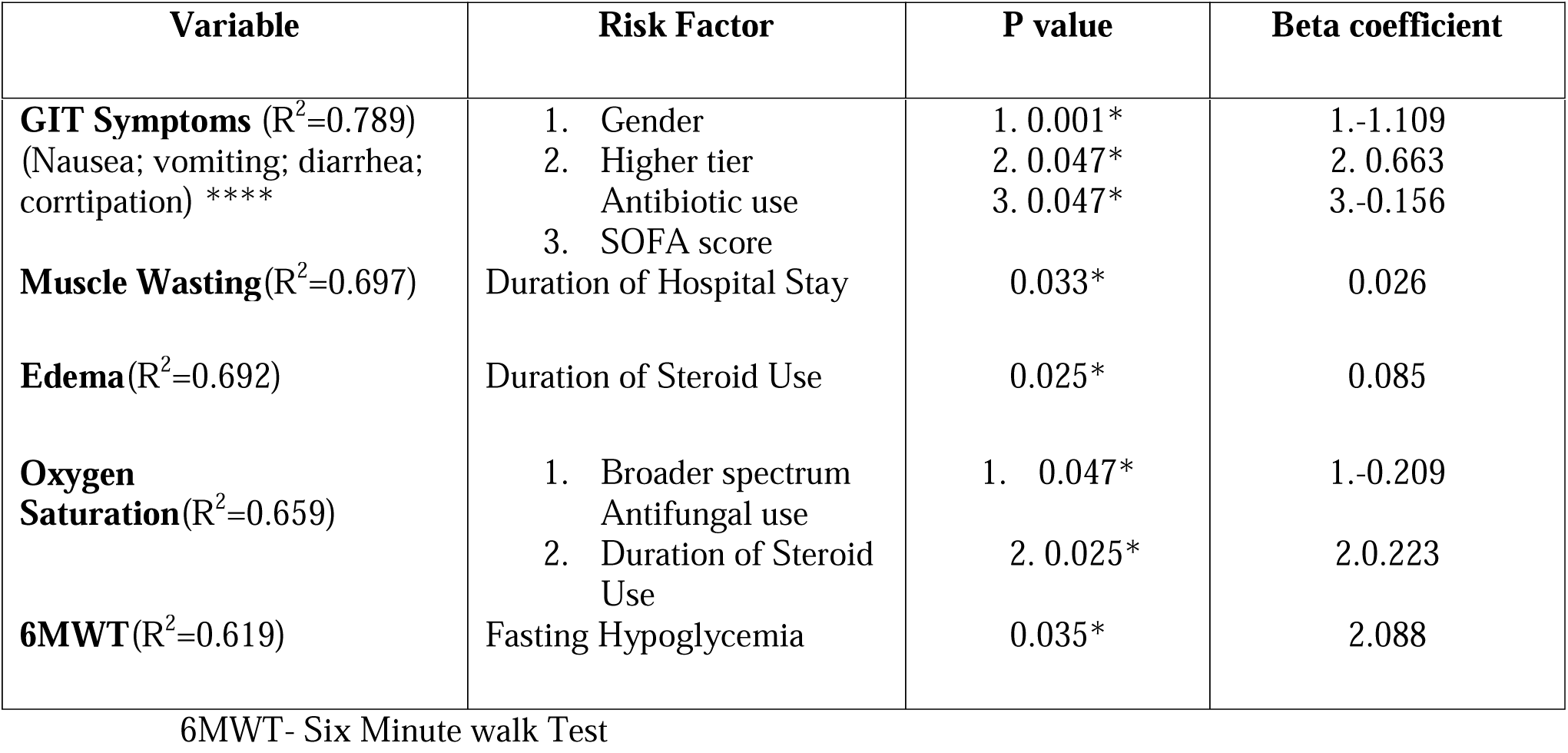
Risk factors for individual variables among baseline characteristics at 0 months.

### At 6 months

Similar analysis at 6 months showed that MOCA score had duration of ICU stay as risk factor. SF-36 domain of PAIN at 6 months had duration of hospital stay as risk factor. Persistent GIT symptoms at 6 months had duration of steroid use as a significant risk factor. Age, Female Gender, Fasting hypoglycemia impacted Single Breath Count. Oxygen saturation at 6 months were influenced by Antifungal use, duration of mechanical ventilation, APACHE 2 score, Fasting Hypoglycemia. 6 Minute Walk Test had Fasting Hypoglycemia as a risk factor. **Table 7**

**Table 7:** Risk factors for individual variables among baseline characteristics at 6 months.

| Variable | Risk Factor | P value | Beta coefficient |
| --- | --- | --- | --- |
| **MOCA score** ($R^2=0.571$) | Duration of ICU Stay | 0.034* | -0.261 |
| **SF 36 (Pain)** ($R^2=0.675$) | Duration of Hospital Stay | 0.023* | -1.376 |
| **GIT Symptoms** ($R^2=0.038$) | Duration of Steroid Use | 0.038* | -0.163 |
| **Single Breath Count**<br>($R^2=0.813$) | 1. Age | 1. 0.031* | 1.-0.190 |
|  | 2. Female Gender | 2. 0.013* | 2. 0.620 |
|  | 3. Fasting Hypoglycemia | 3. 0.005* | 3.1.044 |
| **Oxygen Saturation**<br>($R^2=0.908$) | 1. Broader spectrum Antifungal use | 1. 0.005* | 1.-0.120 |
|  | 2. Duration of Mechanical Ventilation | 2. 0.006* | 2. 0.021 |
|  | 3. APACHE 2 Score | 3. 0.025* | 3. 0.055 |
|  | 4. Fasting Hypoglycemia | 4. 0.002* | 4. 0.833 |
| **6MWT** ($R^2=0.671$) | Fasting Hypoglycemia | 0.007* | 2.277 |

MOCA score-Montreal Cognition Assessment
GIT-Gastrointestinal tract
6MWT- Six Minute walk Test

## DISCUSSION

It was logical in assuming that the Covid 19 pandemic, being a new disease, due to its unique characteristics and high rates of mortality, would impart varied long lasting effects on its survivors. Multiple studies have been conducted in ARDS patients to assess the long-lasting consequences of this disease. Herridge et al in a landmark study concluded that ARDS survivors had long term functional disability after one year discharge from the ICU^(20)^. Needham et al in the ALTOS study, demonstrated impairments in 6MWT and SF-36 outcome measures in ARDS survivors^(31)^.

In the study by Herridge et al, the median age of the ARDS survivors was 45 years and is similar to the characteristic of our study population in which Age at presentation was 50.57. Among our study subjects, 22 were male (73.33%) and 8 were female (26.67%), while in the studies by Herridge et al and Needham et al, there were 56% and 50% males respectively, hence our study was a male skewed population. Of the males, 11 resumed work by 6 months. In the same study by Herridge et al, the patients spent a median of 25 days in the ICU and 48 days in the hospital, median of 21 days of mechanical ventilation was required^(20)^. In our study, the mean duration of hospital stay was 38.47 days, mean duration of ICU stay was 17.73 days, mean duration of mechanical ventilation was 20.30 days. The differences may be attributed to the statistical differences between mean and median. The mean APACHE 2 and SOFA scores in first 24 hours was 11.07 and 5.20 respectively in our study. The study by Herridge et al had a median Apache 2 score of 23. The study by Beigmohammadi et al and Gowda et al established that APACHE 2 and SOFA scores were not accurate predictors of mortality for Covid-19 infected patients^(31,32,33)^. Out of the study subjects, around half of the patients had no prior comorbid illnesses, while the study by Needham et al enrolled all patients with comorbid illnesses among which psychiatric comorbidity and substance abuse were predominant^(31)^. All the study subjects received antivirals with Remdesivir being the predominant one. This is in keeping with fact that the ACTT-1 trial results were released around that time which showed beneficial effects of Remdesivir against Sars-Cov2 ^(34)^. Most of the study subjects were exposed to broad-spectrum high-end antibiotics like Polymyxins, Carbapenems and drugs used for multi drug resistant organisms. This was probably due to prolonged ICU stay, hospital stay, prolonged mechanical ventilation and extended sessions of proning employed which resulted in ventilator associated pneumonia and other sources of sepsis. Among the study subjects, exposure to antifungals was seen in 43%. This is in keeping with fact that a lot of invasive fungal infections were seen in patients infected with Delta Variant of Sars-Cov-2^(35)^. Among the study subjects, 67% of patients received immunomodulator drugs. This was due to the publishing of ACTT-2 trial at that time which brought out the efficacy of immunomodulators like baricitinib^(36)^. Most of the study subjects received 3 and 4 weeks of steroids with the maximum duration being more than 8 weeks. This was in part due to the results of Recovery trial which showed beneficial effects of steroids in Covid-19^(37)^. The studies by Needham et al and Herridge et al had calculated steroid dose equivalent to prednisone given to patients and not duration of steroids. Their studies concluded that steroids were partly responsible for physical impairment by causing muscle weakness upto a period of 6 months^(23)^ ^(31)^.

Most of the patients 60% had CT severity score between 16-20 indicating severe disease^(39,40)^. 36% patients developed sepsis during the hospital stay. The study by Herridge et al and Needham et al do not mention the incidence of in hospital sepsis. Rather illness acquired during ICU stay was mentioned not including hospital acquired illness. Only a small percentage of patients developed stage 3 AKI as per KDIGO. This statistic was similar in number in the study by Herridge et al in the survivor group ^(23)^. During the hospital stay ∼40% patients underwent some form of surgery which included percutaneous tracheostomy, amputation, splenectomy. A slightly larger percentage of survivors underwent tracheostomy among survivors in the study by Herridge et al^(23)^. One patient died after discharge before 6 month follow up in our study.

Mikkelsen at al in his study showed that using a telephone battery was a feasible method to assess neuropsychological functions^(26)^. This was further supported by article by Gordon et al^(27)^. Modrykamien et al in his article described that Montreal Cognitive assessment is one among the standard tools for the evaluation of cognition during follow up visits after ICU discharge^(10)^. Mikkelsen et al concluded that cognitive and psychiatric morbidity are common in survivors of ALI and executive dysfunction was a common morbidity^(26)^. From the baseline characteristics of our study, it is revealed that our subjects had a similar mean age but the duration of ICU stay, duration of mechanical ventilation, degree of hypoxia as per P/F ratios were more in our study subjects which indicates a sicker cohort of patients. These patients were also exposed to a wide variety of antibiotics, antifungals, steroids which were in part due to uncertainty regarding treatment surrounding Sars-Cov-2.

### Cognition

In our study all the patients had a mean MOCA score of 25.17 which signifies mild cognitive dysfunction at discharge and on follow up at 6 months, they had a mean score of 27.07 which signified full recovery after 6 months. This was in contrast to the study by Mikkelsen et al and Rothenhausler et al in which the subjects had long-term cognitive morbidity^(25,26)^.

### Mental Health

Herridge et al in her study used the SF 36 questionnaire to evaluate ARDS patients at follow up ^(23)^. Needham et al in his cohort study found that more than half the survivors of ARDS had substantial symptoms in at least 1 domain of PTSD, depression, general anxiety with frequent co-occurrence. Our study used SF 36 to assess impact of physical conditioning on mental health ^(23,31)^. Mikkelsen et al in his study determined that substantial PTSD, depression, and non-specific anxiety symptoms occurred in survivors of ARDS ^(26)^. Jackson et al found that functional quality of life was less affected in patients who had daily spontaneous awakening and breathing trials at 12 month follow up ^(41)^. In our study, the mental health assessed as per medical outcomes study short form (SF 36) domains revealed that Pain was the only domain that showed good recovery after 6 months. All the other domains were significantly affected at discharge and recovery after 6 months was minimal and barely touched score of 50. Herridge et al in her study showed that the scores for all domains of the SF-36 improved from 3 to 12 months after discharge from the ICU which was in keeping with the findings in our study ^(23)^. However the domain of emotional role limitation had significant recovery in her study which was not reflected in our study^(23)^. Fatigue persisted as a dominantly affected domain suggesting probable neuromuscular side effects of Covid-19. This could mean that Role limitations due to emotions and fatigue could be a significant component of PICS after Sars Cov-2 infection.

### Physical Domains

Those with normal BMI at 0 months and 6 months were different with less number having normal BMI at 6 months. The study by Herridge et al also showed that around 6% change in weight was present in survivors with loss of weight being common^(23)^. In the study by Needham et al, the mean BMI at baseline was 31 and correlated negatively with 6MWT and other physical functions^(31)^. Muscle wasting was seen in majority of our study subjects at discharge which recovered partially or completely on follow up visit at 6 months. This was similar to the study results of Needham et al, Herridge et al where significant decrease in muscle bulk was observed in ARDS survivors. These effects recovered after 6-12 months^(23)(31)^. In our study, some patients had persistent symptoms of nausea, diarrhea, vomiting which recovered by follow up. This was also in keeping with the study by Fan et al in which survivors recovered by 12 months^(38)^. Pedal edema and generalized edema was seen in most of our patients and persisted at 6 months in our study. This was also seen in the study by fan et al Needham et al and Herridge et al ^(23,31,38)^.

Bedside Pulmonary function assessment was assessed by oxygen saturation, Dyspnea scales, single breath count, 6MWT in our study. Oxygen saturation (measured via pulse oximetry) below 88% in room air and requiring domiciliary oxygen support was seen among 20% which improved by 6 months to maintain above 88%. However, most patients with saturation between 88-94% did not show much recovery at 6 months. Dyspnea as assessed by the modified medical research council Dyspnea scale (mMRC), showed that half the patients (56%) had grade 2-3 dyspnea which persisted at the end of 6 months. However, the patients with the severe grade 4 dyspnea, recovered to manageable levels at the end of 6 months. Dyspnea scales, Oxygen saturation were not parameters that were assessed in most ARDS survivors in most other studies probably due to dyspnea scales being subjective and pulse oximeter being considered inaccurate when those studies were published. The Pulse oximeter has only recently gained popularity in recent years as a fifth vital sign^(42)^. The single breath count did not show statistically significant difference in our study subjects but improvement was seen in majority of our patients at 6 months. The 6MWT in our study, revealed significant disability with 40 % having capacities of <250m at discharge. However significant improvement in the same was also seen at follow up at 6 months. The studies by Needham et al, Herridge et al, Fan et al used 6 Minute Walk Test as a chief measure of pulmonary function^(23,31,38)^. In these studies, it was observed that 6 Minute Walk Tests were severely affected at discharge and later improved to some extent at the end of 6 months.

The secondary objective of our study was to elicit risk factors for the systemic syndrome of PICS overall. No baseline characteristic emerged as risk factors when assessed for PICS syndrome as a whole. **Table 4**. This is probably due to the small sample size of the study population. However, the domains when considered individually did reveal some characteristics as risk factors. At ICU discharge or first follow up, higher tier antibiotic use, Female gender, higher SOFA scores appeared to be risk factors for GI symptoms. GI symptoms were not one of the outcomes assessed in the studies by Needham et al and Herridge et al^(23,31)^. Increased duration of hospital stay emerged as a risk factor for muscle wasting. Herridge et al attributed muscle wasting due to a multifactorial cause and partly explained by steroid use and critical illness myopathy^(23,28)^.

Unexpectedly, Broad spectrum antifungal use and longer duration of steroid use was associated with better Oxygen saturation, Fasting hypoglycemia emerged as a risk factor for 6MWT at discharge and follow up. This contrasted with findings by Herridge et al as increased duration of ICU stay, hospital stay, duration of mechanical ventilation, corticosteroid use was associated with lesser distance covered by 6MWT at 3 months^(23)^. This maybe due to the effectiveness of antifungal use and steroids in Sars Cov-2 illness.

At the end of 6 months, some of these effects were no longer seen. Herridge et al also found that effect of steroids did not extend upto 6 months^(23)^. At follow up, duration of hospital stay was a risk factor for cognitive deficits in our study. Other studies did not find significant associations between long-term cognitive impairment and mechanical ventilation, use of sedatives, vasopressors, or analgesic medications; enteral feeding; hypoxia; extracorporeal membrane oxygenation; systolic blood pressure; pulse rate; or length of ICU stay^(43)^. Gastrointestinal symptoms had duration of steroid use as risk factor. Single breath count was affected by older age, female gender, and fasting hypoglycemia. The study by Saleri et al showed a decrease in Single Breath Count among patients recovered from Covid-19 pneumonia ^(44)^. Oxygen saturation was affected by higher APACHE scores, duration of mechanical ventilation, fasting hypoglycemia and type of antifungal use. Broader spectrum of antifungal drug resulted in better oxygen saturation. At the end of 6 months, fasting hypoglycemia persisted as a risk factor for 6MWT. In the study by Herridge et al, Lung injury scores, in hospital illness, use of paralytic agent were risk factors for impaired 6MWT^(23)^

### LIMITATIONS OF THE STUDY

Our study had several limitations. The sample size was not large enough to identify overall risk factors for the Post intensive Care Syndrome as a whole and the study is at risk of being underpowered. Selection bias is a possibility due to the observational nature of the study. Pre registration before data collection could not be done to enhance transparency due to several factors, one among them being the nature of the pandemic situation itself. Psychiatric diagnoses could not be formally established due to absence of structured clinical interviews, but the tools used in our study may serve as a screening tool. The effect of paralytic agents, dose of steroids could not be assessed in our patients.

## CONCLUSION

✓ Sars-Cov-2 virus caused mild cognitive dysfunction at discharge which recovered at 6 months.
✓ It caused long lasting emotional effects and fatigue which may extend beyond 6 months.
✓ It caused muscle wasting and affected pulmonary function of which Oxygen saturation and 6MWT had partial recovery while dyspnea showed full recovery.
✓ Modifiable risk factors exist for each individual aspect of PICS which we can try and affect during in hospital treatment.

Our findings suggest that Post Intensive care Syndrome due to severe Sars-Cov-2 virus affected survivor’s cognitive functions, mental health and caused physical impairment.

Some solutions include:

✓ Critical care Rehabilitation programs involving critical care physicians, psychiatrists, respiratory therapists, occupational therapists and other multidisciplinary teams may help diagnose PICS early and help recovery for life threatening diseases. Such departments exist outside our country but none in our country.
✓ Restricting prolonged steroid use, overaggressive treatment modalities may prevent some aspects of PICS
✓ Fasting hypoglycemia can affect pulmonary function and needs to be addressed.
✓ Fungal infections require aggressive treatment and may have improved outcomes if treated appropriately.
✓ Appropriate Longitudinal studies to identify risk factors in larger cohorts.

## Data Availability

All data produced in the present study are available upon reasonable request to the authors

---

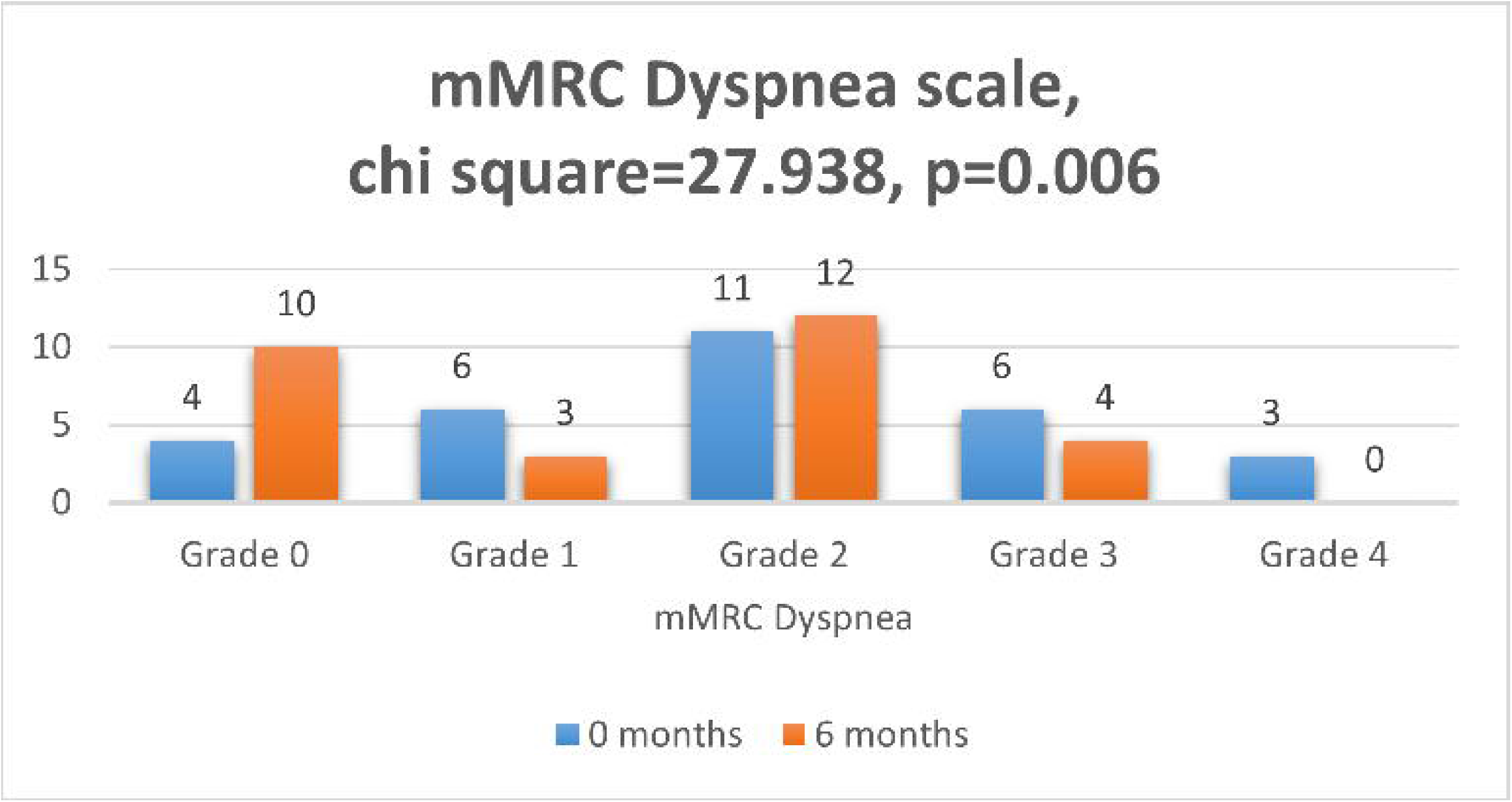

